# Photoacoustic imaging to monitor outcomes during hyperbaric oxygen therapy: Validation in a small cohort and case study in a bilateral chronic ischemic wound

**DOI:** 10.1101/2022.05.12.22274576

**Authors:** Yash Mantri, Aditya Mishra, Caesar A. Anderson, Jesse V. Jokerst

## Abstract

Diseases of the microcirculatory system are well-known risk factors for chronic wound healing. Hyperbaric oxygen therapy (HBO_2_) is a common therapeutic modality that drives oxygen into hypoxic tissue to promote healing. Ischemia/hypoxia are common confounding variables associated with failure of wound progress and/or relapse, and hence it is important to develop tools that map and measure perfusion and oxygen saturation in the wound bed. Photoacoustic (PA) imaging is an ideal tool to address these concerns. Ten patients undergoing HBO_2_ underwent PA oximetry of the left radial artery and forearm pre- and post-HBO_2_; this cohort validated the use of PA imaging in HBO_2_. There was a significant increase in radial artery oxygenation after HBO_2_ (p=0.002) in the validation cohort. PA significantly underestimated arterial oxygenation compared to a pulse oximeter. We also include a case study: a non-diabetic male in his 50s (HB 010) presenting with bilateral ischemic and gangrenous wounds. HB 010 underwent additional scanning of the wound sites both pre- and post-HBO_2_. HB 010 showed higher perfusion and oxygen saturation on the right foot than the left after HBO_2_ which correlated with independent surgical observations. Imaging assisted with limb salvage treatment options by limiting the initial amputation site to only the toes. Hence, this work shows that PA imaging can measure changes in arterial oxygen saturation due to HBO_2_; it can also produce 3D maps of tissue oxygenation and evaluate response to therapy during HBO_2_.

**Key Points:** Photoacoustic oximetry can measure and map changes in arterial oxygenation due to hyperbaric oxygen therapy. Photoacoustic imaging shows changes in perfusion in a patient presenting with bilateral ischemic and gangrenous wounds and thus informing limb salvage treatment.

## Introduction

Diseases of the microcirculatory system (peripheral vascular and arterial disease)(1–3), diabetes(4, 5), proinflammatory vasculitis, and immobility issues(6) are well-known risk factors for chronic wounds.(7, 8) The American Heart Association’s guidelines on managing patients with peripheral artery disease lists non-healing chronic wound formation as a symptom, and recommends that a wound specialist be an integral part of the care team.(9) Chronic wounds have a variety of etiologies and time-courses, but they almost always involve vascular insult and remodeling. Foot ischemia, peripheral neuropathy, and deformities significantly increase the risk of amputation.(10) A study in 2013 evaluating 602 diabetic patients with ischemic foot ulcers treated without revascularization showed that only 50% and 17% healed after minor and major amputations, respectively.(11) Amputation only removes the obvious tissue failure and does not treat the underlying vascular insult.(12) Doctors continue to assess residual tissue survival and amputate more if needed. The goal is to limit amputation.

Common wound treatments work to improve perfusion (how much blood is in tissue) and oxygenation (what percent of the blood is oxygenated). These treatments include sequential compression therapy(13), augmented venous return, and serial debridement(14) to elicit senescent cellular release of growth factors such as VEGF(15) and remove necrotic tissue. More advanced modalities include negative pressure wound therapy (NPWT)(16) and hyperbaric oxygen therapy (HBO_2_).(17) In HBO_2_, pure oxygen is supplied to the patient at elevated pressure to promote blood and tissue oxygenation. HBO_2_ can stimulate vasculogenic stem cells to initiate both angiogenesis and neovascularization of the wound bed.(18) HBO_2_ improves wound perfusion and can thus promote wound healing.(17)

Multiple studies have shown that the use of HBO_2_ significantly reduces wound recurrence (0-15%) due to the higher oxygen supply to healing tissue.(19–21) Perfusion and oxygenation are also linked to recurrence. A meta-analysis of 19 studies presented in the *NEJM* showed that roughly 40% of patients have recurrence within one year after healing and 60% within three years.(22) Ischemia/hypoxia associated with PAD(23, 24) are some of the most common confounding variables for wound relapse.(25–27) Reduced perfusion of the skin and soft tissues results in localized ischemia and necrosis leading to ulceration.(1) A retrospective study on 189 limbs with critical limb ischemia showed that patients with successful revascularization had significantly higher limb salvage rates than partial/failed revascularization.(28) Clearly, there is a rigorous body of research showing that restoration of adequate microvascular perfusion and tissue oxygenation is essential to ensure low recurrence rates while also supporting more complete wound healing.(29)

Transcutaneous oximetry (TCOM) is one approach to measuring oxygenation,(30) but it is an ensemble approach that is surface-weighted and reports a global average value across the estimated wound area (commonly including healthy tissue)—it cannot discriminate between different areas of the wound nor map perfusion and oxygenation levels in three dimensions. TCOM values are also quite variable and highly operator dependent even in healthy subjects(31); thus clear diagnostic discriminators have yet to be developed. Diffused optical imaging, Laser Doppler imaging, near infrared spectroscopy (NIRS), and others have also been proposed but can only study the first few mm of tissue(32–36). Thus, visual inspection still remains the most common approach to monitoring wounds including response to therapy during HBO_2_.(37)

Previously, we and others have reported the use of photoacoustic (PA) imaging to monitor changes in peripheral perfusion, oxygenation, and angiogenesis in chronic wounds.(37–39) Photoacoustic imaging is a hybrid imaging technique that combines optical spectroscopy with ultrasound. PA leverages optical absorbers such as melanin in skin and hemoglobin in blood to generate endogenous contrast. The PA signal is overlayed on conventional ultrasound (US) images to provide spatio-functional imaging.(40) We and others have shown that PA signal can report hemoglobin oxygenation.(41–45) This is logical because oxyhemoglobin and deoxyhemoglobin absorb at different wavelengths (**Fig. 1**). Beyond oxygenation, PA signal can also correspond to perfusion based on total signal (i.e., more total signal = more total hemoglobin);(46) concurrently, ratiometric signal can be used to quantify oxygenation.(42)

**Figure 1.**
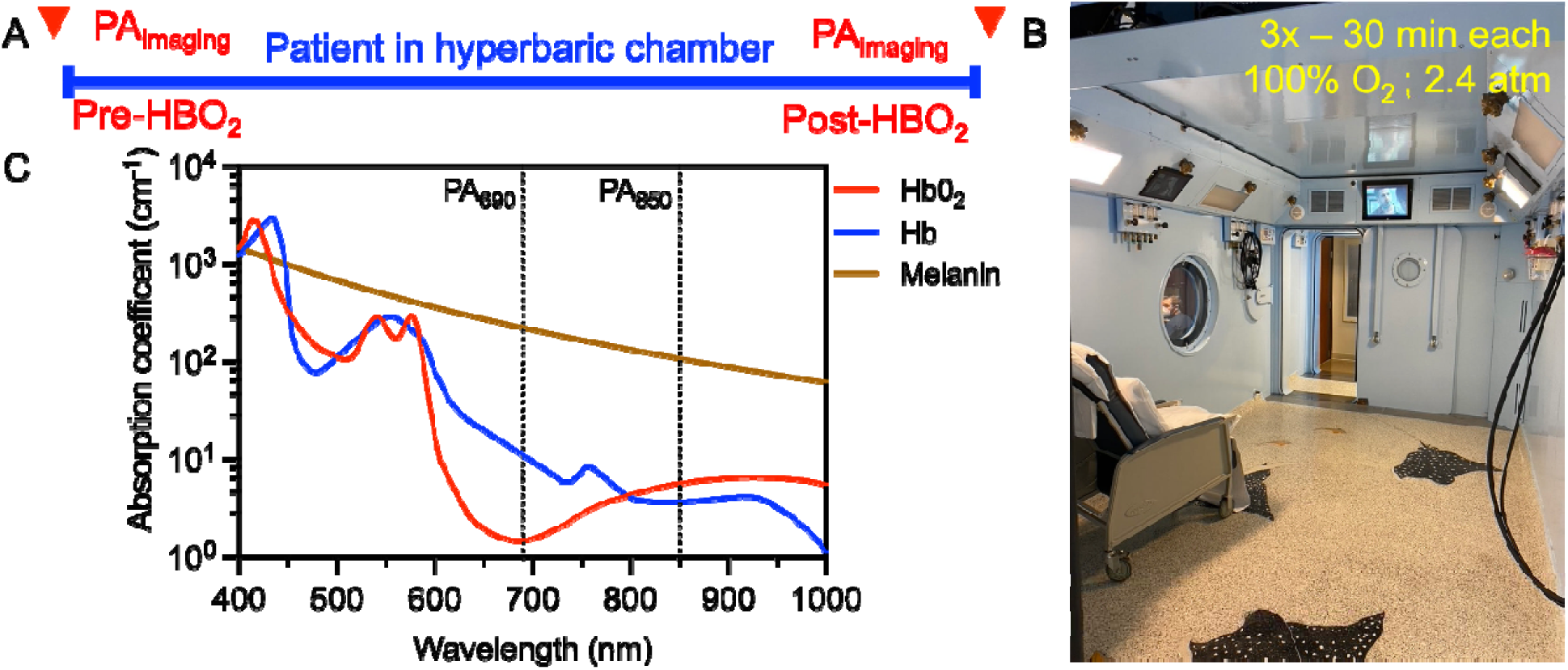
Study design. **A**. All participants were scanned twice: (i) pre and (ii) post hyperbaric oxygen therapy (HBO_2_). The scans took place within five minutes of entering or exiting the chamber. **B**. The multiplace HBO_2_ chamber at UCSD seats 6 patients at a time. All participants underwent the same HBO_2_ treatment: A descent to 45 FSW (Foot Sea Water; ∼2.4 atm absolute pressure), 100% oxygen delivered via a hood over three 30-min periods with two 5-min air breaks. **C**. The absorption coefficient of the major optical absorbers (oxy/deoxyhemoglobin and melanin) in tissue. Photoacoustic imaging was performed at two wavelengths (850 and 690 nm) (47),(48).

In this work, we present the use of PA imaging to map changes in tissue oxygenation in a chronic lower limb ischemic wound due to HBO_2_. We first validated the use of PA imaging on HBO_2_ patients without wounds. We then hypothesize a significant increase in arterial oxygen saturation after HBO_2_ therapy. We next imaged a patient with a chronic ischemic lower limb wound showing areas of good, average, and poor perfusion in healthy, wound edge, and wound area respectively. This study illustrates the value of PA imaging in mapping perfusion in the wound bed in three dimensions.

## Methods

### Participants

This work was compliant with the ethical guidelines for human experimentation as stated in the 1975 Deceleration of Helsinki. The University of California San Diego’s Human Research Protections Program approved this study (IRB# 191998). Written informed consent was obtained from all subjects before participation. Inclusion criteria were: (i) ≥18 years old and ability to provide consent and (ii) undergoing HBO_2_ and/or presenting with a chronic wound. Exclusion criteria were: (i) presence of bloodborne pathogens and (ii) presence of implants in the imaging region. A total of ten participants were recruited consecutively for this study at the UCSD hyperbaric Medicine and Wound Care Center, Encinitas, CA. PA imaging was performed on all participants. We also used an over-the-counter finger pulse oximeter (ZAcurate, Texas, USA) as an independent measurement. One subject (HB 009) was dropped from the study due to corrupted file storage resulting in a loss of data for the post-HBO_2_ scan. Subjects HB001-08 had no visible wounds and were treated for complications related to radiation exposure for cancer treatment (**Table S1**). HB001-08 served as controls to validate the PA imaging technique. HB010 was a unique case presenting with variable levels of ischemic insult. He had dry gangrene of all toes with tissue viability more established at the proximal ankle. Hypoxic tissue between the proximal ankle and gangrenous toes were considered threatened with unclear perfusion status. Hence, HB010 served as the primary subject for this case study.

### Hyperbaric Oxygen Therapy

All participants were treated in a multiplace hyperbaric chamber and underwent the same HBO_2_ treatment protocol. All patients remained in a seated position for descent to 45 FSW (Foot Sea Water; ∼2.4 atm absolute pressure) without any incident. 100% oxygen was delivered via a hood with three 30-min periods and two 5-min air breaks. All patients ascended without any incidents.

### Case history

Patient (HB 010) presented with multiple upper and lower extremity ischemic insults with gangrenous toes and fingers. Wounds were at variable stages with more viable tissue proximal compared to dried gangrenous tissue more distal. The skin surface of all treated areas were dark in color and cool to touch. The distal tip of multiple fingers on both hands also exhibited frank necrosis and gangrene. Before presenting at the UCSD Hyperbaric Medicine and Wound Care Center, Encinitas, CA, he had already tolerated 6 HBO_2_ cycles without complications such as claustrophobia or otic barotrauma. Before PA scanning, patient had undergone an additional 19 HBO_2_ sessions. He was able to tolerate treatment well without incident.

After PA scanning, the patient underwent thorough debridement of all necrotic and infected tissue as well as amputation of all gangrenous and necrotic toes in both feet. The right foot was treated first just after the PA scan followed by the left foot the following day. The ischemic fingertips remained stable with dry gangrene and were considered more salvageable than the feet. Operative debridement was therefore postponed for a later date.

### Photoacoustic (PA) imaging

All PA imaging was done using a commercially-available LED-based system (AcousticX from Cyberdyne Inc., Tsukuba, Japan).(49) We used two multiwavelength LED arrays operating at 850 and 690 nm, pulse width 70 ns, and 4 KHz repetition rate. The AcousticX uses a 128-element US transducer operating at a central frequency of 7 MHz, 0.38 × 6 cm field of view, and a bandwidth of 80.9%.(46) All scans were done using a hand-held transducer. All participants were scanned twice within 5 min of entering or existing the HBO_2_ chamber (**Fig. 1A**).

We used a proprietary gel pad from Cyberdyne Inc. and 1 cm thick layer of sterile US gel to couple the transducer with the skin surface. We also used a sterile US transducer cover (CIV-Flex, product no. 921191, AliMed Inc., Dedham, MA) for all scans to prevent cross contamination.

We scanned the left forearm and radial artery of all patients due to a lack of curvature, relative hairlessness, and ease of scanning in that region. A PA scan was done on the forearm, which measured approximately 4 cm x 4 cm for 3D mapping of the vasculature in that region. The radial artery served as a site for arterial oxygen saturation measurements. The transducer was not moved while measuring oxygen saturation in the radial artery.

The left forearm and left radial artery of HB010 served as control areas. Five imaging sites were identified on each foot: (i) the proximal-most superior healthy tissue, (ii) expected survival area, (iii) threatened area, (iv) plantar foot, and (v) heel. These areas were identified by the attending wound care physician (C.A.A.) as (i) healthy tissue proximal to the wound site, (ii) region that appeared adequately perfused and hence expected to survive, (iii) region showing signs of ischemia and threatened to become necrotic with high potential amputation risk, (iv) plantar foot region with necrotic tissue in need of extensive debridement, and (v) heel with necrotic tissue also in need of extensive debridement, respectively (**Fig. 2**). Before PA scanning, we first removed all wound dressings and cleaned the wound site with sterile isotonic saline. The five imaging spots on both feet were scanned as shown in **Fig. 2**.

**Figure 2.**
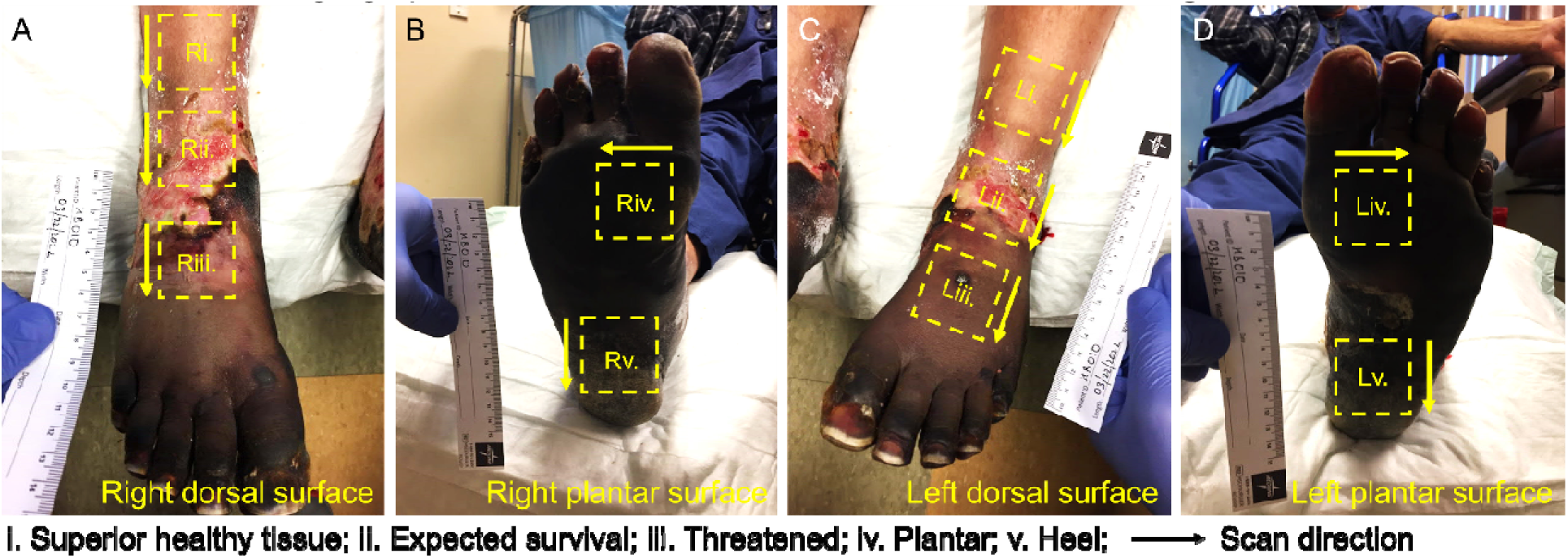
Case presentation HB010. A male patient presents following vasopressor-induced ischemic insults to both lower and upper extremity resulting in gangrene of all toes and multiple finger tips on both hands. The patient (HB 010) received 25 HBO_2_ sessions prior to enrolling in this study. Five discrete imaging spots were identified based on the expected survivability of tissue in consultation with the attending physician. (i) healthy tissue superior to the wound, (ii) region that appeared adequately perfused and hence expected to survive, (iii) threatened region with signs of ischemic injury at elevated amputation risk, (iv) plantar region with frank necrosis in need of extensive debridement, and (v) heel with frank necrosis also in need of extensive debridement. The arrows denote the scan direction.

### Image Processing

All images were visualized and exported using proprietary software built for the LED-PA acquisition system (Cyberdyne Inc. version 2.00.10). A minimum of 70 frames of 8-bit PA and US images were exported for each scan. PA data contained the frames were acquired sequentially at 690 and 850 nm. Frames were analyzed using region of interest (ROI)-based analysis (ImageJ, version 2.1.0/1.53c, and Fiji extension) using a previously reported method.(42) 3D maximum intensity projections (MIP) were used to visualize the imaging regions in three dimensions.

The relative concentrations of oxy-and deoxy hemoglobin (HbOxy and HbR, respectively) can be calculated by solving **Equation 1**. Here, *ε*_*HbR*_ (*λ*_*i*_) and *ε*_*HbOxy*_ (λ _*i*_) are the known molar extinction coefficients (cm^−1^M^−1^) of HbR and HbOxy at wavelengths of λ_*i*_ (850 and 690 nm), respectively. ROIs annotating the radial artery were custom-drawn for each scan. We measured the mean PA intensity within the ROI at 850 and 690 nm. The extinction coefficient for HbR and HbOxy were preset into the system by the manufacturer. We used the same preset extinction coefficients for total HbR and HbOxy calculations. The total oxygen saturation (sO_2_) was calculated using **Equation 2**.

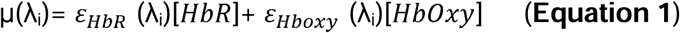

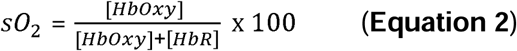

### Statistics

We measured changes in PA oximetry due to HBO_2_. We used a paired, two-tailed t-test to compare radial artery sO_2_ pre-and-post HBO_2_. We used a Bland-Altman analysis to compare PA measured radial artery sO_2_ and pulse oximetry measured sO_2_. Based on previously reported values in literature, a mean bias above 4% would be considered clinically significant.(50)

## Results and Discussion

### Validating photoacoustic oximetry for HBO_2_ patients (HB 001-008)

We scanned the left volar forearm and left radial artery of all participants. All participants underwent two scans: (i) pre-and (ii) post-HBO_2_ treatment. Both scans were within five minutes of entering and exiting the chamber. We also used a commercially available pulse oximeter as an independent sO_2_ measurement. Pulse oximeter readings showed a significant increase (p = 0.008, paired t-test) in sO_2_ after HBO_2_ treatment (**Table S1**). PA imaging of the radial artery also showed a similar trend with a paired, two-tailed t-test showing a significant increase in arterial sO_2_ (s_a_O_2_) after HBO_2_ treatment (p = 0.002, **Fig. 3C**). While pulse oximetry is a peripheral endpoint measurement, PA imaging can map the vasculature in the forearm in 3D showing increasing oxygen saturation after HBO_2_ (**Fig. 3D-E**). Bland-Altman analysis showed that PA imaging significantly underestimates oxygen saturation (mean difference = −6.48%, **Fig. S1**). Others have shown that sO_2_ measured via a pulse oximeter can significantly differ from arterial blood gas (ABG) measurements.(51) ABG is the gold standard technique to measure oxygen and carbon dioxide in the blood stream, but it is an invasive measurement and was hence avoided in this study.(52) A study in 102 critically ill patients showed that an pulse oximeter reading of above 94% was necessary to ensure arterial sO_2_ above 90% as measured using ABG.(50)

**Figure 3.**
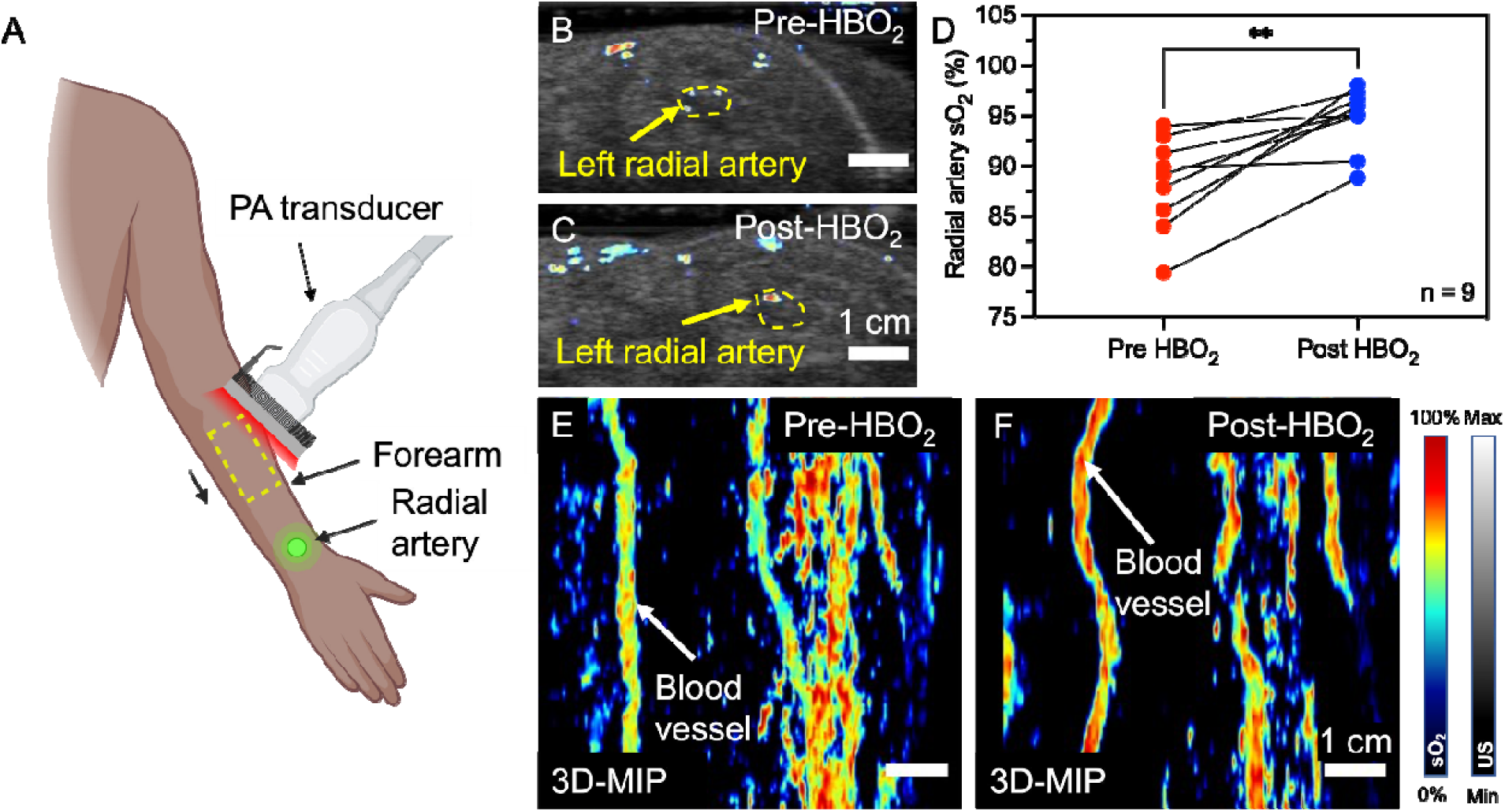
Photoacoustic imaging of the left radial artery and volar forearm. **A**. For validation, we scanned the left radial artery and volar forearm before and after HBO_2_ treatment. **B-C**. Axial cross-section of the left radial artery pre and post HBO_2_ respectively. Yellow dotted region and arrow mark the radial artery. **C**. PA oximetry of the radial artery shows a significant increase (p = 0.002) in arterial oxygenation after HBO_2_ treatment. A minimum of 50 frames were quantified for each data point. **D**. 3D maximum intensity projections (MIP) viewed from the top show an increase in blood oxygen saturation after HBO_2_.

Skin thickness and skin tone (melanin concentration) can have a major impact on optical oximetry measurements. Increasing skin thickness and darker skin tones can significantly reduce light penetration into tissue, thus resulting in erroneous measurements. We have previously reported a pilot study with tools to compensate for the differences in skin tone.(41) This previous work suggested that subjects with darker skin tones show increased light absorption on the skin surface resulting in lower s_a_O_2_ readings, reduced penetration depth, and obscuring underlying vasculature. Although the participants enrolled in this study had relatively lighter skin tones (Fitzpatrick range 1-3, **Table S1**), the differences in skin tone could be a reason why our data did not match the pulse oximeter. This could be further compounded by the thicker layer of skin on the wrist compared to the fingertip used for pulse oximetry. A more extensive study with more subjects is underway to better understand this phenomenon.

### Case study: Severe bilateral ischemic insults (HB 010)

HB 010: An otherwise healthy male in his 50s presented with gangrene of all toes. He received 25 HBO_2_ sessions prior to PA scanning for this study. Pulse oximetry measurements were considerably limited secondary to the presence of gangrenous fingertips. PA oximetry of the radial artery, however, was able to capture measurements demonstrating significant increase in oxygen saturation (7.8% increase in s_a_O_2_ after HBO_2_; **Fig. S2**) and blood perfusion (more PA signal, **Supplementary Videos 1 and 2**). The radial artery with intact healthy skin served as a key control for our imaging technique.

PA oximetry of the right and left dorsal foot surface showed a higher oxygen saturation (more red pixels) on the right foot compared to the left (**Fig. 4**). PA imaging after HBO_2_ treatment further confirmed higher perfusion levels in the right foot (**Fig. 5**). Post-HBO_2_ of the Rii region (expected survival, **Fig. 5B and E**) showed the highest increase in oxygen saturation and perfusion, thus suggesting a better prognosis and higher chance of salvageability. The same region on the left foot did not exhibit any changes in the PA signal suggesting less salvageability potential (**Fig. 5H and K**).

**Figure 4.**
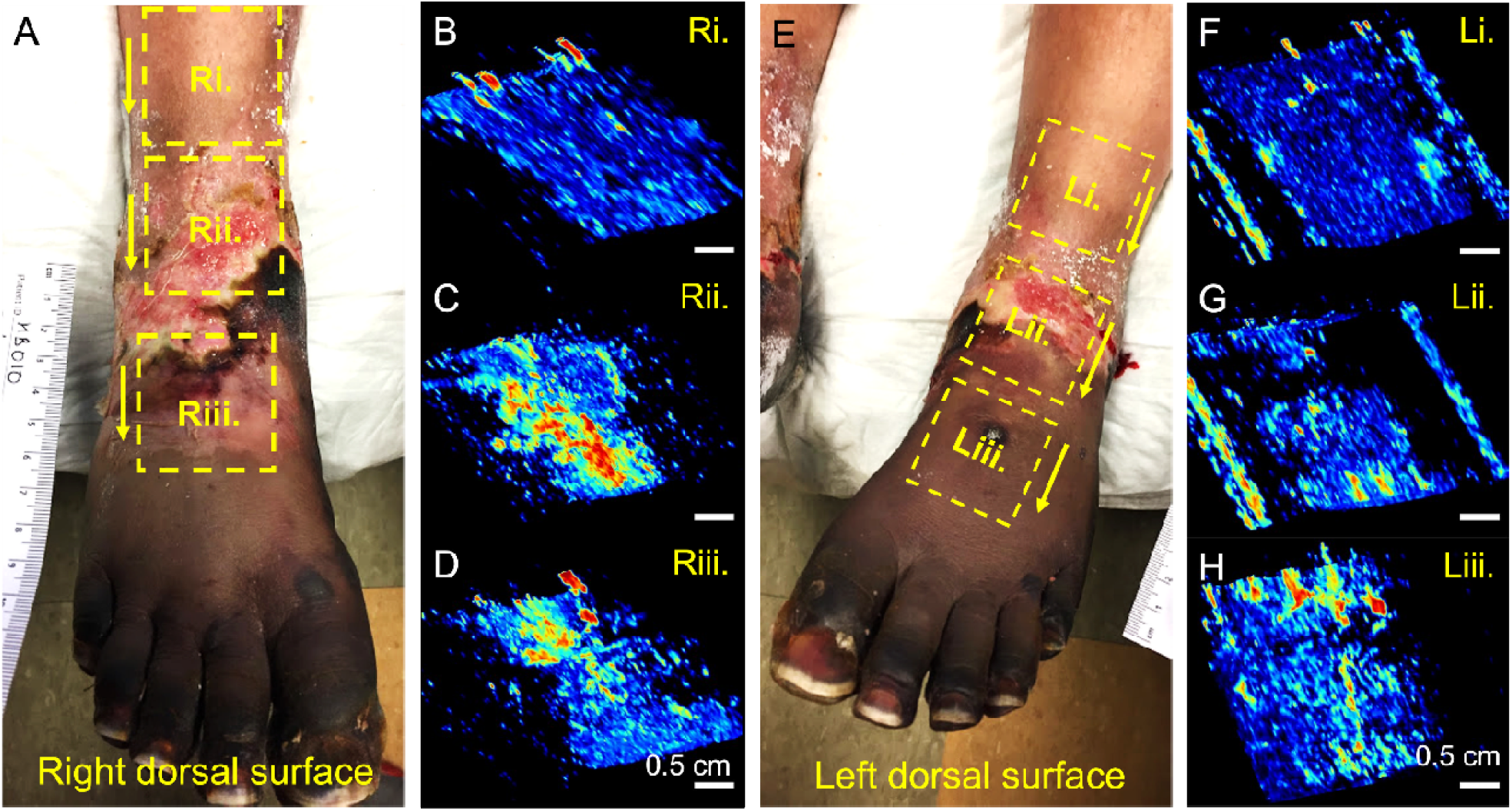
PA oximetry of the right (A-D) and left (E-H) dorsal surface pre-HBO_2_. Three regions were scanned on the dorsal surface on each foot. (i) Superior heathy region, (ii) expected survival, and (iii) threatened region. **B-D and F-H** 3D-MIP images of the right and left foot respectively. More red pixels indicate higher oxygen saturation. The right foot has more red pixels, thus indicating better tissue oxygenation in these regions (Rii (**C**) vs Lii (**G**)). Scale bars in the PA images represent 0.5 cm, and the yellow arrows in the photographs indicate the scan direction.

**Figure 5.**
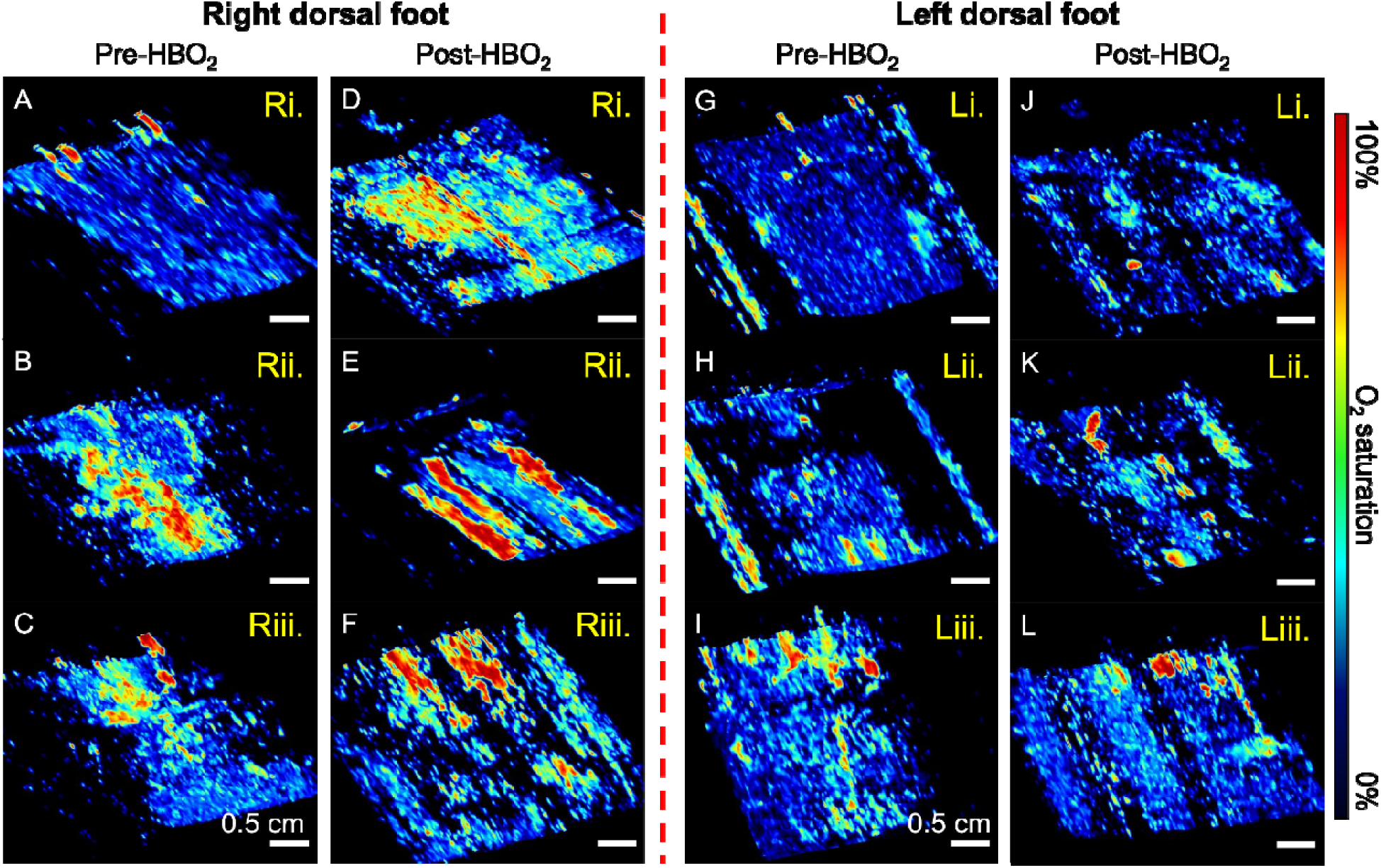
PA oximetry of the dorsal foot surface pre-and post-HBO_2_ shows bette perfusion on the right foot. **A-C** and **D-F** show 3D-MIP images of the right dorsal foot surface pre-and post-HBO_2_, respectively. All three regions (Ri, Rii, and Riii) show more red pixels, thus indicating higher oxygen saturation after HBO_2_ treatment. **G-I** and **J-L** show 3D-MIP images of the left dorsal foot pre-and post-HBO_2_, respectively. Unlike the right foot, the left foot showed no significant increase in oxygen saturation perhaps indicating reduced perfusion leading to a worse prognosis for the left foot compared to the right foot. All scale bars represent 0.5 cm. All scans were reconstructed using a minimum of 750 B-mode frames.

The main advantage of using PA imaging is the ability to 3D map blood perfusion and oxygen saturation—both are key parameters for wound healing.(38, 53) The 3D MIP images showed some increase in perfusion and oxygenation up to the Riii (**Fig. 5C and F**). This informed therapy by limiting the initial amputation to the toes instead of the entire foot (**Fig. S3**). Moreover, PA was able to demonstrate objective levels of perfusion/oxygenation which can provide the surgeon with additional clinical information which may translate into potentially salvageable tissue. Tissue without perfusion could be excised earlier minimizing infection risk and morbidity. An ideal study would have longitudinally monitored this patient using PA imaging. Our previous work showed that PA monitoring of angiogenesis can predict wound healing.(37) Unfortunately, there is an increased risk of infection and secondary trauma after amputation and extensive debridement (**Fig. S3 and S4**); hence, subsequent imaging was avoided.

To validate PA imaging as a diagnostic tool for a wound patient undergoing HBO_2_, one potential ideal case would be a unilateral ischemic wound to compare between healthy and threatened ischemic tissue for the same patient. One limitation of this study is the lack of a healthy baseline measurement of foot perfusion and oxygen saturation. Although we see a clear increase in perfusion and oxygenation after HBO_2_, we cannot conclude if the change were sufficient to predict wound healing. We note that this case was extremely challenging with many confounding variables affecting the healing process. Some of these include but are not limited to a history of secondary trauma related to inadequate off loading and occasionally poor patient compliance.

PA imaging of the plantar surface did not reveal any changes in perfusion due to HBO_2_ treatment. The plantar surface had thick necrotic tissue with darkened eschar (blackened, **Fig. 2B and D**). Hence, the imaging reveals no conclusive evidence of an increase in blood perfusion or oxygen saturation in the plantar region.

Compared to other techniques such as TCOM, toe, and ankle brachial index currently employed in predicting wound healing, PA offers the ability to map the wound bed with high spatiotemporal resolution overlayed with functional information about perfusion and oxygen saturation.(37–39) TCOM suffers from long acquisition times and low inter user reliability.(54) Ankle and toe brachial index has shown promise but it is limited to peripheral perfusion measurements whereas PA imaging can be performed anywhere.(55) Diffused optical and multispectral imaging are also upcoming techniques used to predict wound amputation but this light in-light out approach has limited depth penetration (mm scale).(35, 36, 56) The light in-sound out approach of PA imaging allows deeper (cm scale) penetration into tissue. Our previous work on predicting wound healing showed that PA imaging could predict wound therapy outcomes better than conventionally used techniques (AUC = 0.915).(37)

### Limitations

In this work, we used LED-based PA imaging for ease of use, cost effectiveness, and aid clinical translation. The LEDs we used operate at 1000-fold lower fluence compared to conventionally used OPO-lasers.(49) The use of high-powered OPO-lasers would allow for more light to be delivered to underlying tissue and increase imaging sensitivity.(57) But the laser-based systems are significantly more expensive and sometimes operate above the maximum permissible exposure limit set by ANSI.(46, 58)

## Conclusions

This preliminary study explores the use of photoacoustic (PA) imaging to monitor changes in blood perfusion and oxygen saturation in patients undergoing hyperbaric oxygen therapy (HBO_2_). The validation study in nine HBO_2_ patients scanned pre-and post-HBO_2_ showed a significant (p = 0.002) increase in oxygen saturation due to HBO2. We also present a challenging case of subject with bilateral ischemic wounds also undergoing HBO_2_. 3D PA oximetry mapping showed that the right foot was better perfused and oxygenated after HBO_2_. The imaging informed the amputation procedure by initially limiting amputation to the toes. The main finding of this work is that PA imaging can measure changes in arterial oxygen saturation and produce 3D maps of the oxygen distribution in threatened tissue.

## Data Availability

All data produced in the present study are available upon reasonable request to the authors

## Acknowledgements

Figure 1A and 3A were made using BioRender.com. We acknowledge help from all the nurses at the Hyperbaric Medicine and Wound healing Center, University of California San Diego, Encinitas, CA. We also thank all study participants. This work was supported by the National Institutes of Health under Grant R21 AG065776; Internal funds from the University of California San Diego, under the Galvanizing Engineering in Medicine Program.

## Data Availability

All raw data is available upon reasonable request from the corresponding author.

## References

1. Singer AJ, Tassiopoulos A, Kirsner RS. Evaluation and management of lower-extremity ulcers. New England Journal of Medicine 2017;377(16):1559–1567.

2. Farber A. Chronic limb-threatening ischemia. New England Journal of Medicine 2018;379(2):171–180.

3. Boyko EJ, Monteiro-Soares M, Wheeler SG. Peripheral arterial disease, foot ulcers, lower extremity amputations, and diabetes. Diabetes in America 3rd edition 2018.

4. Blakytny R, Jude E. The molecular biology of chronic wounds and delayed healing in diabetes. Diabetic Medicine 2006;23(6):594–608.

5. Okonkwo UA, DiPietro LA. Diabetes and wound angiogenesis. International journal of molecular sciences 2017;18(7):1419.

6. Raeder K, Jachan DE, Müller-Werdan U, Lahmann NA. Prevalence and risk factors of chronic wounds in nursing homes in Germany: A Cross-Sectional Study. International Wound Journal 2020;17(5):1128–1134.

7. Alam W, Hasson J, Reed M. Clinical approach to chronic wound management in older adults. Journal of the American Geriatrics Society 2021;69(8):2327–2334.

8. FrykbergRobert G. Challenges in the treatment of chronic wounds. Advances in wound care 2015.

9. Gerhard-Herman MD, Gornik HL, Barrett C, Barshes NR, Corriere MA, Drachman DE, Fleisher LA, Fowkes FGR, Hamburg NM, Kinlay S. 2016 AHA/ACC guideline on the management of patients with lower extremity peripheral artery disease: a report of the American College of Cardiology/American Heart Association Task Force on Clinical Practice Guidelines. Journal of the American College of Cardiology 2017;69(11):e71–e126.

10. Mills Sr JL, Conte MS, Armstrong DG, Pomposelli FB, Schanzer A, Sidawy AN, Andros G, Committee SfVSLEG. The society for vascular surgery lower extremity threatened limb classification system: risk stratification based on wound, ischemia, and foot infection (WIfI). Journal of vascular surgery 2014;59(1):220–234. e222.

11. Elgzyri T, Larsson J, Thörne J, Eriksson K-F, Apelqvist J. Outcome of ischemic foot ulcer in diabetic patients who had no invasive vascular intervention. European Journal of Vascular and Endovascular Surgery 2013;46(1):110–117.

12. Forsythe RO, Apelqvist J, Boyko EJ, Fitridge R, Hong JP, Katsanos K, Mills JL, Nikol S, Reekers J, Venermo M. Performance of prognostic markers in the prediction of wound healing or amputation among patients with foot ulcers in diabetes: A systematic review. Diabetes/Metabolism Research and Reviews 2020;36:e3278.

13. Lurie F, Bittar S, Kasper G. Optimal compression therapy and wound care for venous ulcers. Surgical Clinics 2018;98(2):349–360.

14. Manna B, Nahirniak P, Morrison CA. Wound debridement. 2018.

15. Dixon D, Edmonds M. Managing diabetic foot ulcers: pharmacotherapy for wound healing. Drugs 2021;81(1):29–56.

16. Huang C, Leavitt T, Bayer LR, Orgill DP. Effect of negative pressure wound therapy on wound healing. Current problems in surgery 2014;51(7):301–331.

17. Thackham JA, McElwain DS, Long RJ. The use of hyperbaric oxygen therapy to treat chronic wounds: a review. Wound Repair and Regeneration 2008;16(3):321–330.

18. Lam G, Fontaine R, Ross FL, Chiu ES. Hyperbaric oxygen therapy: exploring the clinical evidence. Advances in skin & wound care 2017;30(4):181–190.

19. Goldman RJ. Hyperbaric oxygen therapy for wound healing and limb salvage: a systematic review. PM&R 2009;1(5):471–489.

20. Basile C, Montanaro A, Masi M, Pati G, De Maio P, Gismondi A. Hyperbaric oxygen therapy for calcific uremic arteriolopathy: a case series. Journal of nephrology 2002;15(6):676–680.

21. Davis J, Heckman J, DeLee J, Buckwold F. Chronic non-hematogenous osteomyelitis treated with adjuvant hyperbaric oxygen. J Bone Joint Surg Am 1986;68(8):1210–1217.

22. Armstrong DG, Boulton AJ, Bus SA. Diabetic foot ulcers and their recurrence. New England Journal of Medicine 2017;376(24):2367–2375.

23. Bus SA. Priorities in offloading the diabetic foot. Diabetes/metabolism research and reviews 2012;28:54–59.

24. Okazaki J, Matsuda D, Tanaka K, Ishida M, Kuma S, Morisaki K, Furuyama T, Maehara Y. Analysis of wound healing time and wound-free period as outcomes after surgical and endovascular revascularization for critical lower limb ischemia. Journal of vascular surgery 2018;67(3):817–825.

25. Strauss MB, Miller SS, Aksenov IV. Challenges of wound healing. Challenges of 2011.

26. Hess CT, Kirsner RS. Orchestrating wound healing: assessing and preparing the wound bed. Advances in skin & Wound care 2003;16(5):246–257.

27. Chase SK, Melloni M, Savage A. A forever healing: the lived experience of venous ulcer disease. Journal of vascular nursing 1997;15(2):73–78.

28. Bae J-I, Won JH, Han SH, Lim SH, Hong YS, Kim J-Y, Kim JD, Kim J-S. Endovascular revascularization for patients with critical limb ischemia: impact on wound healing and long term clinical results in 189 limbs. Korean Journal of Radiology 2013;14(3):430–438.

29. Kobayashi N, Hirano K, Nakano M, Ito Y, Ishimori H, Yamawaki M, Tsukahara R, Muramatsu T. Prognosis of critical limb ischemia patients with tissue loss after achievement of complete wound healing by endovascular therapy. Journal of vascular surgery 2015;61(4):951–959.

30. Arsenault KA, McDonald J, Devereaux P, Thorlund K, Tittley JG, Whitlock RP. The use of transcutaneous oximetry to predict complications of chronic wound healing: A systematic review and meta-analysis. Wound Repair and Regeneration 2011;19(6):657–663.

31. Blake DF, Young DA, Brown LH. Transcutaneous oximetry: variability in normal values for the upper and lower limb. Diving and hyperbaric medicine 2018;48(1):2.

32. Lentsch GR, Mobasher P, Mizzoni C, Koenig K, Tromberg B, Georgakoudi I, Ganesan A, Balu M. In vivo optical imaging of vitiligo skin grafting treatment using multiphoton microscopy and reflectance confocal microscopy (Conference Presentation). Photonics in Dermatology and Plastic Surgery 2020: International Society for Optics and Photonics, 2020; p. 112–1100.

33. Holland A, Martin H, Cass D. Laser Doppler imaging prediction of burn wound outcome in children. Burns 2002;28(1):11–17.

34. Patry J, Laurencelle L, Bélisle J, Beaumier M. Vascular Assessment in Patients With a Lower Limb Wound: A Correlational Study of Photoplethysmography and Laser Doppler Flowmetry Toe Pressure Techniques. Journal of diabetes science and technology 2020:1932296820979973.

35. Maheshwari N, Marone A, Altoé M, Kim H, Bajakian D, Hielscher A. Pilot study on monitoring ulcer healing with diffuse optical imaging in a patient cohort affected by peripheral arterial disease (PAD). Optical Diagnostics and Sensing XXII: Toward Point-of-Care Diagnostics: SPIE, 2022; p. 47–53.

36. Khalil MA, Kim HK, Kim I-K, Flexman M, Dayal R, Shrikhande G, Hielscher AH. Dynamic diffuse optical tomography imaging of peripheral arterial disease. Biomedical optics express 2012;3(9):2288–2298.

37. Mantri Y, Tsujimoto J, Donovan B, Fernandes CC, Garimella PS, Penny WF, Anderson CA, Jokerst JV. Photoacoustic monitoring of angiogenesis predicts response to therapy in healing wounds. Wound Repair and Regeneration 2021.

38. Wang Y, Zhan Y, Harris LM, Khan S, Xia J. A portable three-dimensional photoacoustic tomography system for imaging of chronic foot ulcers. Quantitative imaging in medicine and surgery 2019;9(5):799.

39. Mantri Y, Tsujimoto J, Penny WF, Garimella PS, Anderson CA, Jokerst JV. Point-of-Care Ultrasound as a Tool to Assess Wound Size and Tissue Regeneration after Skin Grafting. Ultrasound in Medicine & Biology 2021;47(9):2550–2559.

40. Hariri A, Moore C, Mantri Y, Jokerst JV. Photoacoustic imaging as a tool for assessing hair follicular organization. Sensors 2020;20(20):5848.

41. Mantri Y, Jokerst JV. Impact of skin tone on photoacoustic oximetry and tools to minimize bias. Biomedical Optics Express 2022;13(2):875–887.

42. Hariri A, Wang J, Kim Y, Jhunjhunwala A, Chao DL, Jokerst JV. In vivo photoacoustic imaging of chorioretinal oxygen gradients. Journal of biomedical optics 2018;23(3):036005.

43. Zhang HF, Maslov K, Sivaramakrishnan M, Stoica G, Wang LV. Imaging of hemoglobin oxygen saturation variations in single vessels in vivo using photoacoustic microscopy. Applied physics letters 2007;90(5):053901.

44. Bulsink R, Kuniyil Ajith Singh M, Xavierselvan M, Mallidi S, Steenbergen W, Francis KJ. Oxygen saturation imaging using LED-based photoacoustic system. Sensors 2021;21(1):283.

45. Wang Y, Hu S, Maslov K, Zhang Y, Xia Y, Wang LV. In vivo integrated photoacoustic and confocal microscopy of hemoglobin oxygen saturation and oxygen partial pressure. Optics letters 2011;36(7):1029–1031.

46. Mantri Y, Dorobek TR, Tsujimoto J, Penny WF, Garimella PS, Jokerst JV. Monitoring peripheral hemodynamic response to changes in blood pressure via photoacoustic imaging. Photoacoustics 2022:100345.

47. Prahl S. Optical absorption of hemoglobin. http://omlcogiedu/spectra/hemoglobin 1999.

48. Jacques SL. Melanosome absorption coefficient. Oregon Medical Laser Center 1998;7.

49. Hariri A, Lemaster J, Wang J, Jeevarathinam AS, Chao DL, Jokerst JV. The characterization of an economic and portable LED-based photoacoustic imaging system to facilitate molecular imaging. Photoacoustics 2018;9:10–20.

50. Louw A, Cracco C, Cerf C, Harf A, Duvaldestin P, Lemaire F, Brochard L. Accuracy of pulse oximetry in the intensive care unit. Intensive care medicine 2001;27(10):1606–1613.

51. Sinex JE. Pulse oximetry: principles and limitations. The American journal of emergency medicine 1999;17(1):59–66.

52. Castro D, Patil SM, Keenaghan M. Arterial blood gas. StatPearls [Internet]: StatPearls Publishing, 2021.

53. Tonnesen MG, Feng X, Clark RA. Angiogenesis in wound healing. Journal of Investigative Dermatology Symposium Proceedings: Elsevier, 2000; p. 40–46.

54. Ercengiz A, Mutluoglu M. Hyperbaric Transcutaneous Oximetry. 2017.

55. Reed GW, Young L, Bagh I, Maier M, Shishehbor MH. Hemodynamic assessment before and after endovascular therapy for critical limb ischemia and association with clinical outcomes. JACC: Cardiovascular Interventions 2017;10(23):2451–2457.

56. Squiers JJ, Thatcher JE, Bastawros DS, Applewhite AJ, Baxter RD, Yi F, Quan P, Yu S, DiMaio JM, Gable DR. Machine learning analysis of multispectral imaging and clinical risk factors to predict amputation wound healing. Journal of Vascular Surgery 2022;75(1):279–285.

57. Erfanzadeh M, Zhu Q. Photoacoustic imaging with low-cost sources; A review. Photoacoustics 2019;14:1–11.

58. Delori FC, Webb RH, Sliney DH. Maximum permissible exposures for ocular safety (ANSI 2000), with emphasis on ophthalmic devices. JOSA A 2007;24(5):1250–1265.

