## Supplementary material for "Photoacoustic imaging to monitor outcomes during hyperbaric oxygen therapy: Validation in a small cohort and case study in a bilateral chronic ischemic wound": SM1

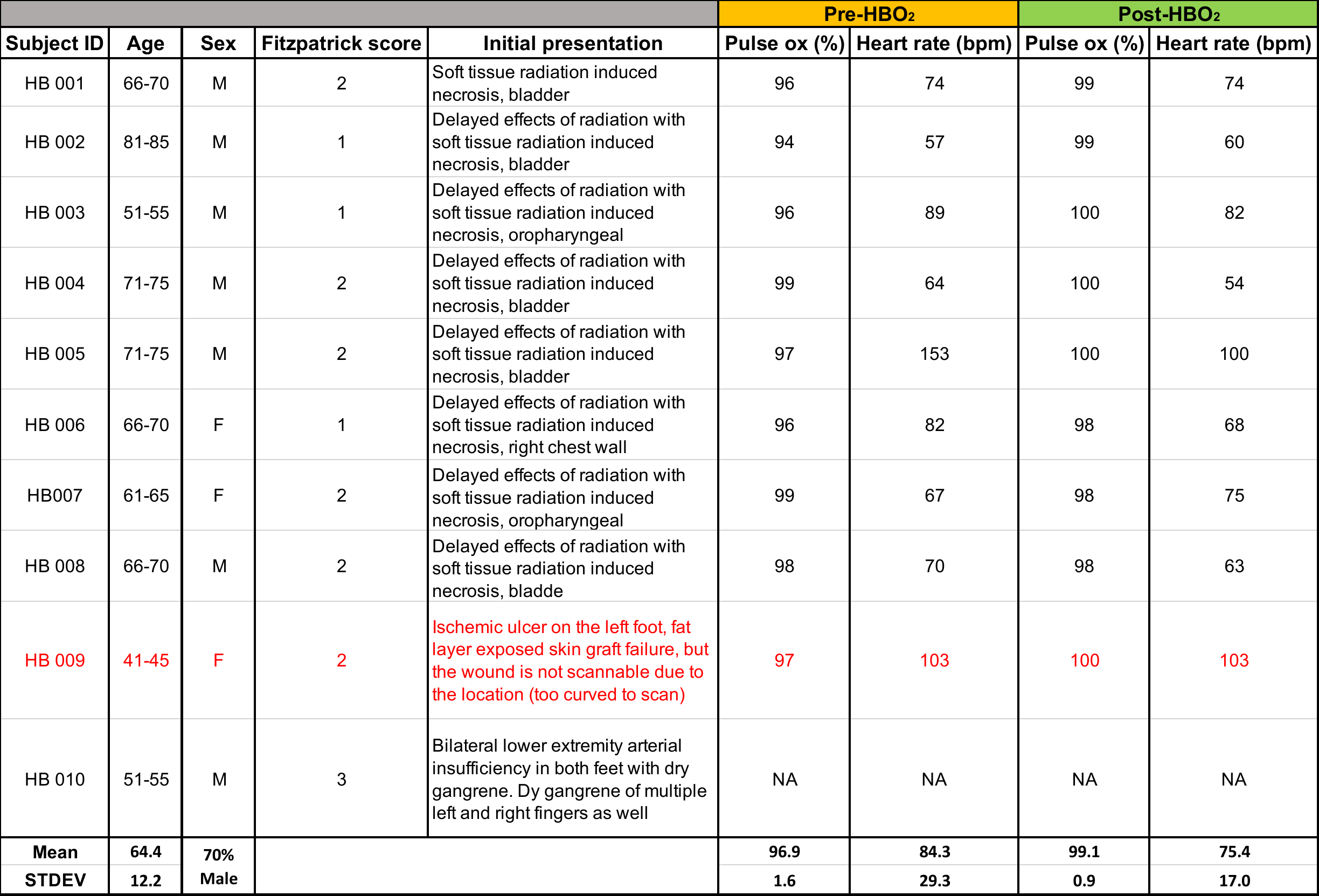

**Table S1.** **Participant demographics.** HB 001-008 served as controls to validate the use of PA imaging for HBO_2_ patients. HB 009 was dropped from analysis due to an error in file storage. HB 010 was a unique case presenting with bilateral ischemic wounds.

MATLAB script:

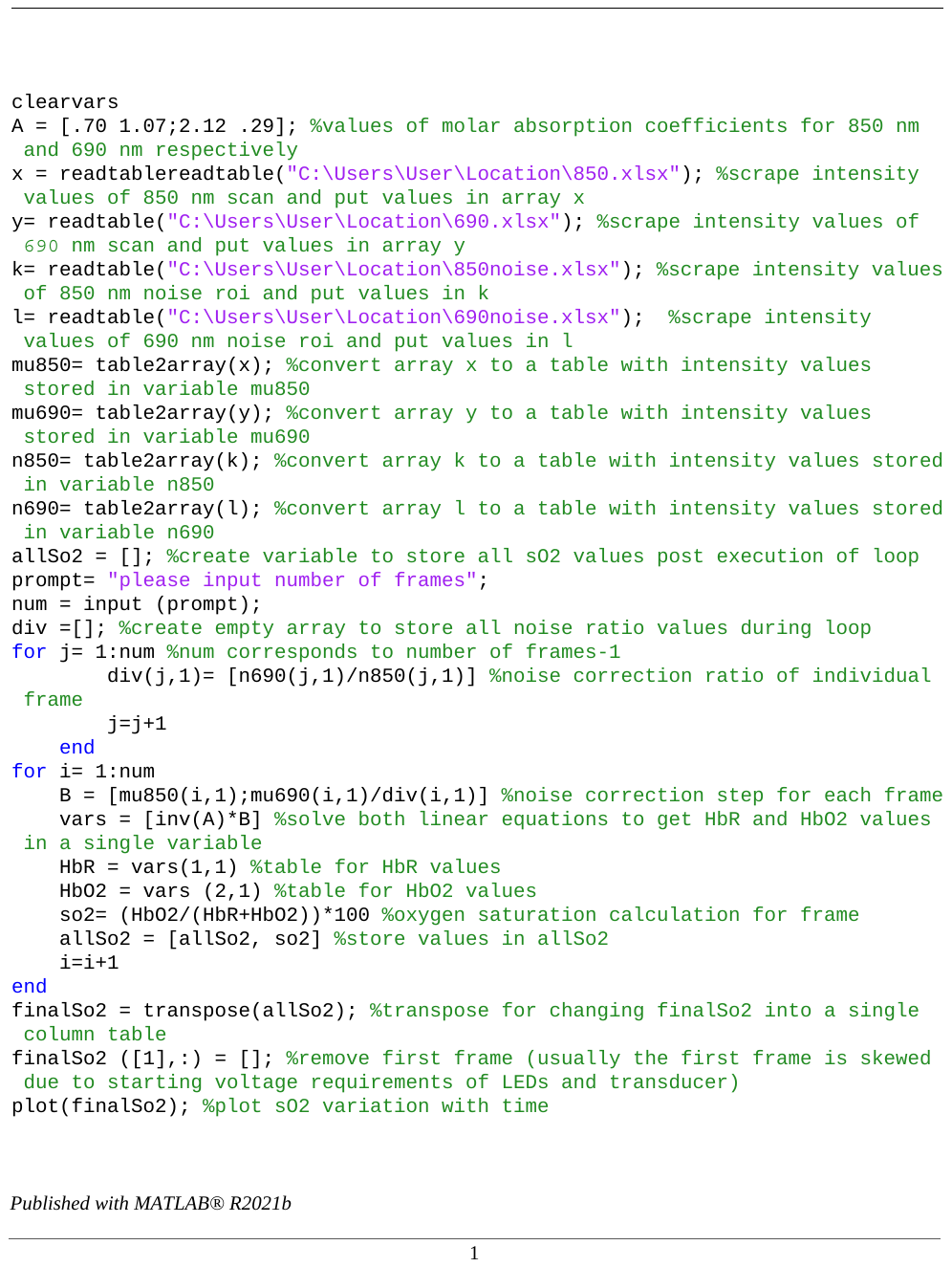

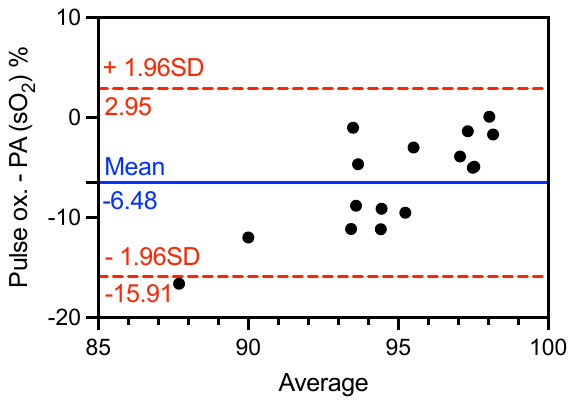

**Figure S1**. Bland-Altman analysis of oxygen saturation measured using a pulse oximeter and PA imaging.

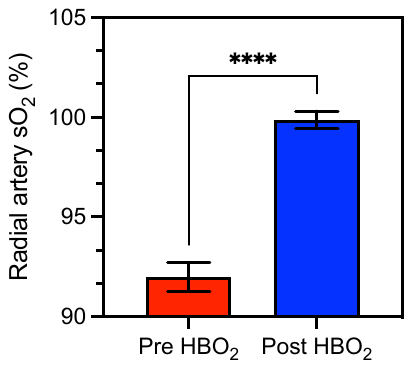

**Figure S2.** Pre- and post-HBOT radial artery oxygen saturation for HB 010 measured using PA oximetry shows a significant increase (p<0.0001) in arterial oxygenation after HBOT.

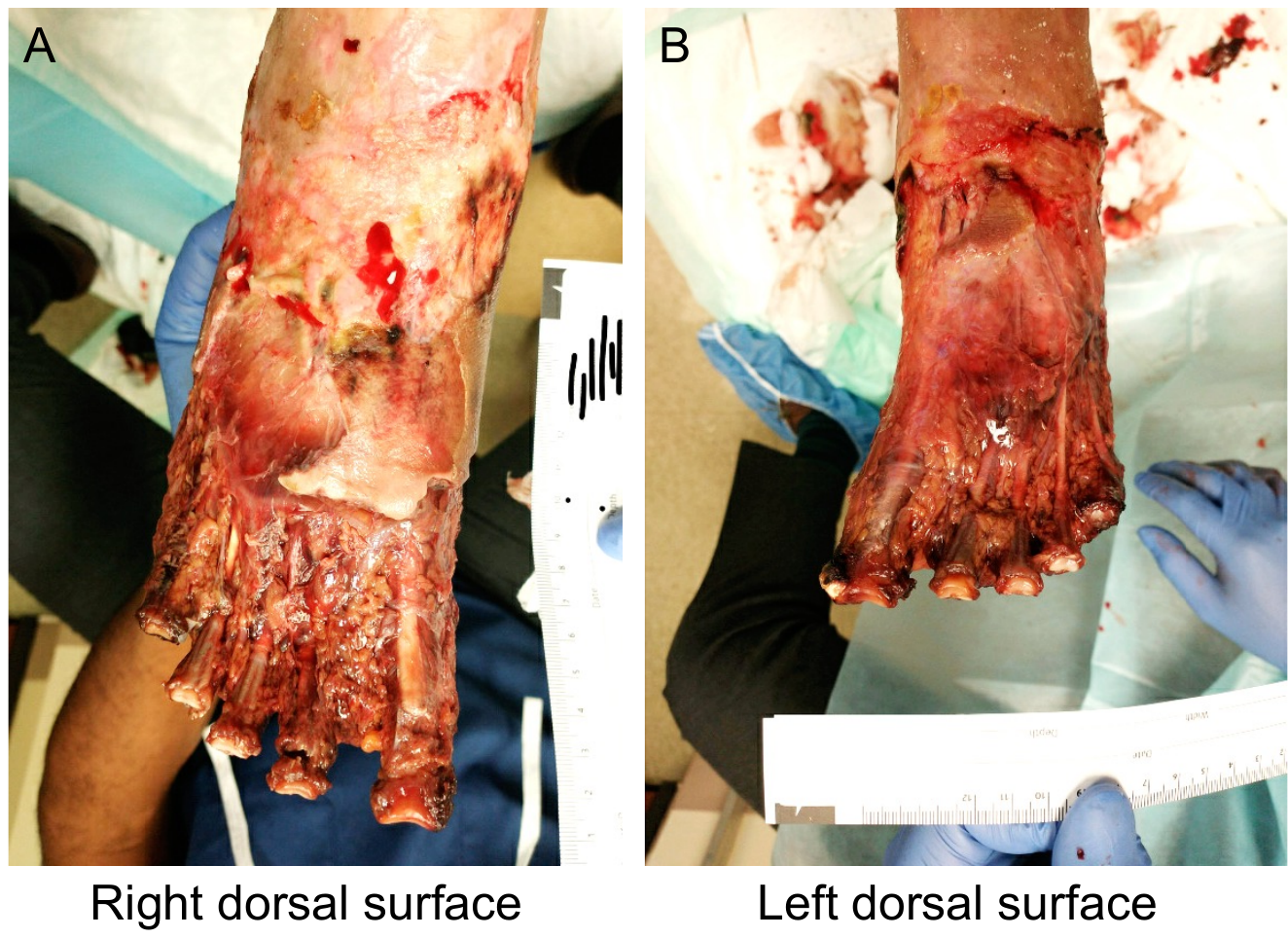

**Figure S3**. Post toe amputation images of the right and left feet (dorsal surface).

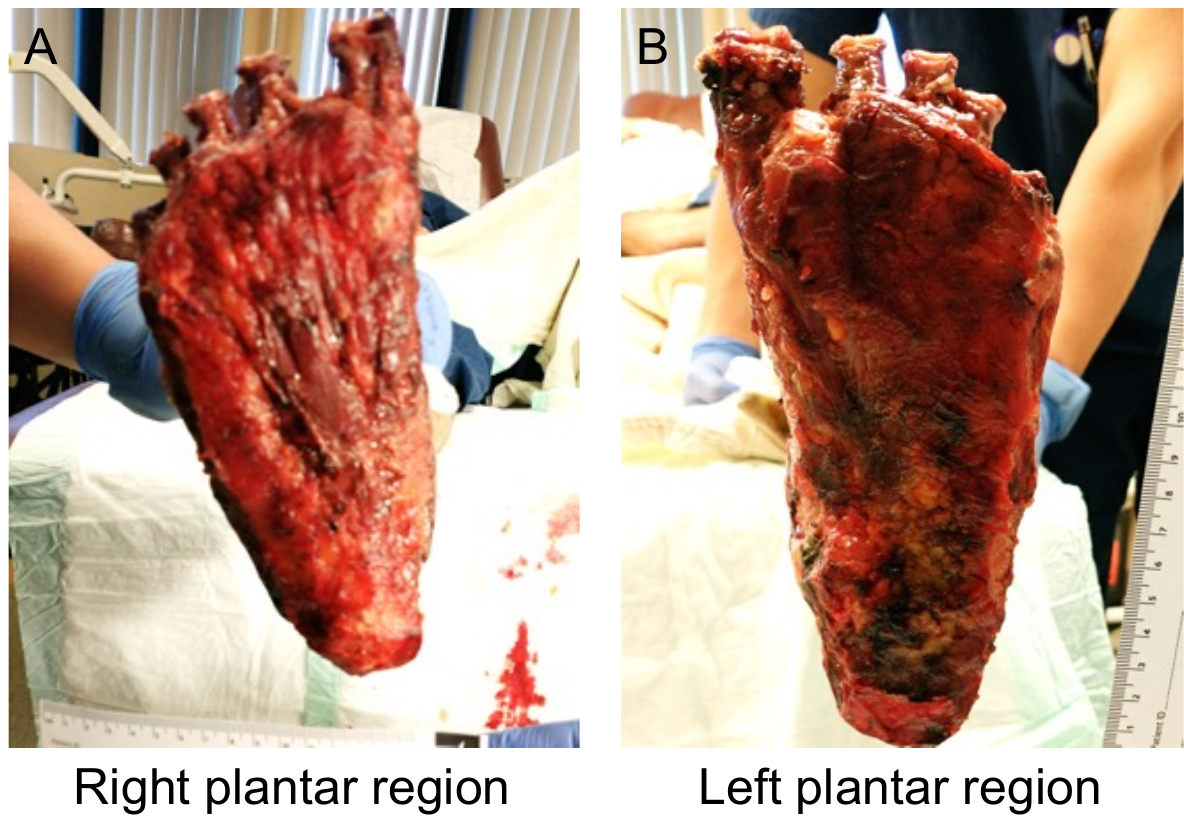

**Figure S4**. Post toe amputation images of the right and left feet (plantar region).

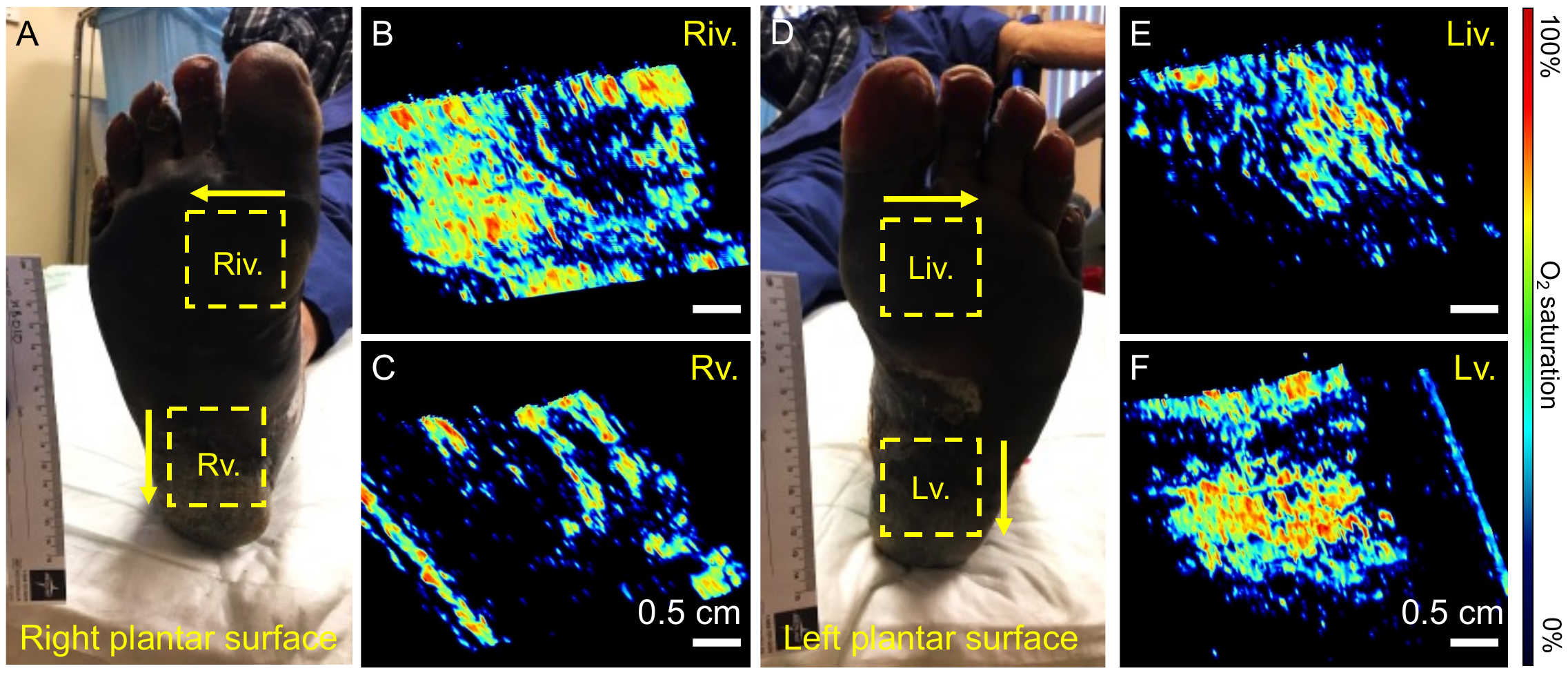

**Figure S5. PA oximetry of the right (A-C) and left (D-F) plantar region pre-HBO_2_.** No conclusive differences were seen between the two feet. Yellow arrows indicate scan direction.

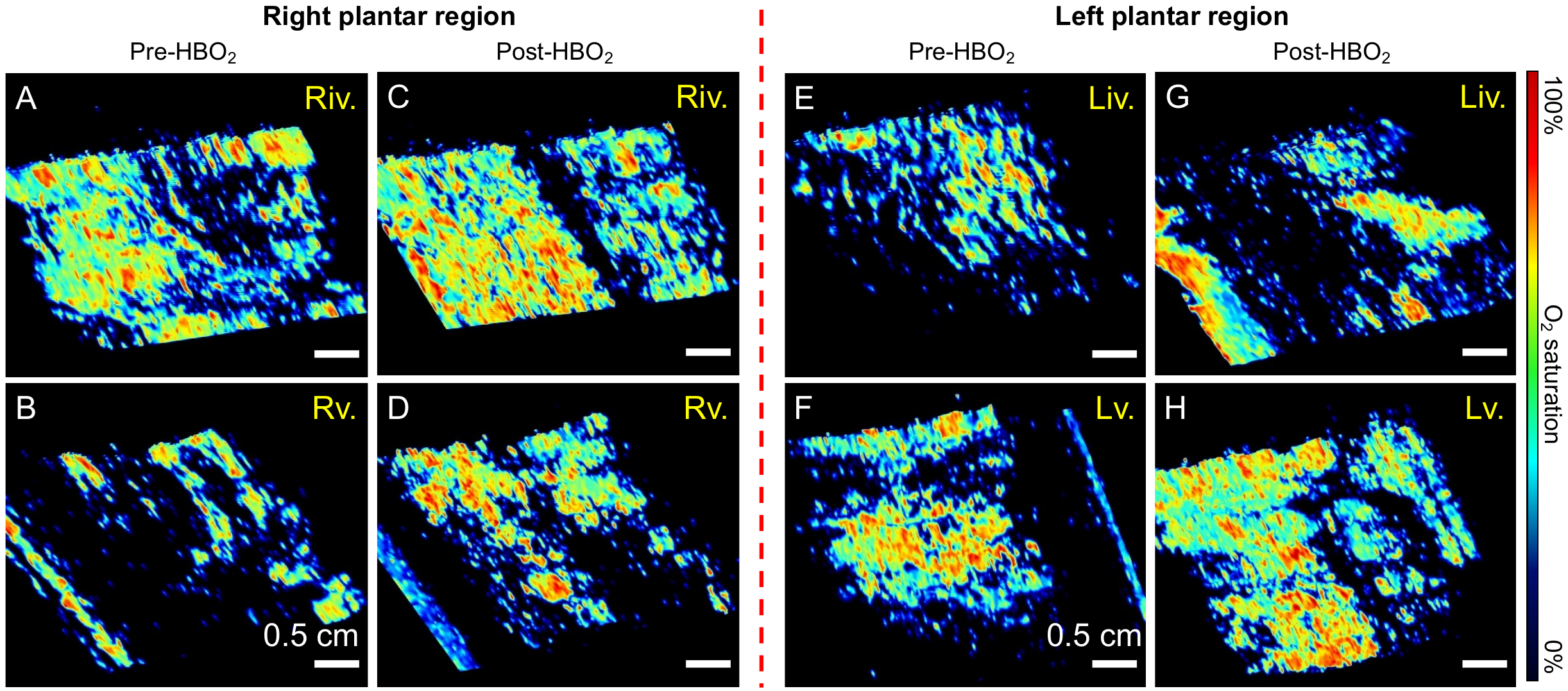

**Figure S6. PA oximetry of the right (A-D) and left (E-H) plantar region pre- and post-HBO_2_ respectively.** No PA differences were seen between the two feet.
